# Discovering Subtypes with Imaging Signatures in the Motoric Cognitive Risk Syndrome Consortium using Weakly-Supervised Clustering

**DOI:** 10.1101/2024.10.11.24315328

**Authors:** Bhargav Teja Nallapu, Ali Ezzati, Helena M. Blumen, Kellen K. Petersen, Richard B. Lipton, Emmeline Ayers, V G Pradeep Kumar, Srikanth Velandai, Richard Beare, Olivier Beauchet, Takehiko Doi, Hiroyuki Shimada, Sofiya Milman, Sandra Aleksic, Joe Verghese

## Abstract

**INTRODUCTION:** Understanding the heterogeneity of brain structure in individuals with the Motoric Cognitive Risk Syndrome (MCR) may improve the current risk assessments of dementia.

**METHODS:** We used data from 6 cohorts from the *MCR consortium* (N=1987). A weakly-supervised clustering algorithm called HYDRA was applied to volumetric MRI measures to identify distinct subgroups in the population with gait speeds lower than one standard deviation (1SD) above mean.

**RESULTS:** Three subgroups (Groups A, B & C) were identified through MRI-based clustering with significant differences in regional brain volumes, gait speeds, and performance on Trail Making (Part-B) and Free and Cued Selective Reminding Tests.

**DISCUSSION:** Based on structural MRI, our results reflect heterogeneity in the population with moderate and slow gait, including those with MCR. Such a data-driven approach could help pave new pathways toward dementia at-risk stratification and have implications for precision health for patients.

## 1. BACKGROUND

Dementia is marked by a progressive decline in cognitive and functional. Accessible and informative clinical indicators that help identify individuals at risk of incident dementia could enable targeted, earlier and more effective interventions. Like Mild Cognitive Impairment (MCI), Motoric Cognitive Risk (MCR) syndrome is a pre-dementia state characterized by subjective cognitive complaints coupled with objectively measured impairment.^1^ While MCI which is characterized by reduced performance on objective cognitive tests, MCR is characterized by slow gait speed and subjective cognitive impairment ^2^. Prior studies indicate that MCR is a significant risk factor for dementia in older adults^3^. Compared to MCI, MCR does not require neuropsychological assessments, which are generally time-consuming. This makes MCR a more efficient and accessible solution for risk stratification of dementia in community-dwelling adults and across various settings, from primary care to specialized neurology clinics ^4^. Both components of MCR, slow gait and subjective cognitive impairment, are also independently predictive of future cognitive decline and incident dementia – but the MCR phenotype has higher predictive validity for dementia than either of the individual components alone ^5,6^.

In older adults, slow gait speed can stem from various factors such as neurological issues, muscle-related conditions, arthritis, or a combination of these factors. Neurodegeneration caused by various dementia etiologies, such as Alzheimer’s disease (AD), is considered one of the primary reasons behind declines in both gait speed and cognitive function^7^. The brain regions that are known to mediate executive functions (EFs), such as the frontal and prefrontal-lobe networks, also control gait ability^8,9^. These brain regions are involved in integrating information from many cortical sensory systems, modulate and produce goal-directed actions and behavior ^10^. Atrophy in these regions causes both cognitive and gait decline, concurrently with the aging process ^11-14^. Preliminary intervention trials using cognitive training or brain stimulation to enhance EF have also shown improvements in gait velocity ^15,16^. MCR, despite these shared neuroanatomical pathways, describes people who are still cognitively intact but with cognitive complaints and slowing of gait. The precise neuroanatomical signatures that correspond to the clinical manifestations in MCR, however, are unknown. To study the biological underpinnings of MCR, an MCR-neuroimaging consortium was established, incorporating data from seven cohorts across five different countries, and three continents. that collected structural MRIs, gait speeds, cognitive assessments, and other clinical symptoms. This consortium provides a valuable opportunity to study neurodegeneration patterns as identified through structural MRIs and to connect these findings with MCR, its components, and other clinical symptoms.

There is substantial neuroanatomical heterogeneity in preclinical stages of dementia ^17^. Several studies have linked MCR to specific patterns of brain atrophy, specifically in prefrontal, supplementary motor, insular and motor cortices ^13,14^. However, very few data-driven studies have explored the variability of neuroanatomical patterns in MCR and its components^14^. Therefore, in this study, we adopted a machine learning based data-driven approach to investigate structural brain differences among participants in the MCR consortium using volumetric imaging. We employed a novel semi-supervised clustering approach called HYDRA (Heterogeneity through Discriminative Analysis) ^18^, to identify subgroups with distinct structural brain patterns within the MCR Consortium. Several other approaches were previously proposed to reveal the inherent disease heterogeneity. But most of these methods either relied on predefined clinical subgroups, ignoring multivariate relationships in the data ^19,20^, or applied clustering directly to brain anatomies, risking the identification of normal inter-individual variability, some of which could be due to confounding factors such as sex, age etc. rather than disease-specific heterogeneity ^21,22^. The HYDRA method enables us to mitigate these challenges and disentangles the heterogeneity in a population, using another reference population. This was achieved by leveraging data from a reference group composed of individuals with faster gait speeds (population FG, those with gait speed faster than 1 standard deviation above the population mean within each cohort). Identifying subgroups with homogenous neuroanatomical patterns in a cohort population with gait variability has a potential to further our understanding of biological underpinnings of MCR, and in developing interventions for dementia.

## 2. Methods

### 2.1. Participants

Data from 2,007 older adults in the MCR consortium was examined. The data were obtained from 7 different cohorts and 5 different countries. The cohorts were : (i) the Central Control of Mobility in Aging Study (CCMA) ^7,23^ in the US, (ii) the LonGenity study ^24^ in the US, (iii) the Einstein Aging Study (EAS) ^2,25^ in the US, (iv) the Tasmanian Study of Cognition and Gait (TASCOG) ^26,27^ in Australia, (v) the National Center for Geriatrics and Gerontology–Study of Geriatric Syndromes (NCGG-SGS) ^28,29^ in Japan, (vi) the Kerala-Einstein Study ^30^ in India, and (vii) the Gait and Alzheimer’s Interactions Tracking study (GAIT) ^9,31^ in France. All the study procedures in each of the cohorts were approved by the local institutional review boards. All the cohorts excluded individuals with prevalent dementia.

Data from all the cohorts were used in the analyses. We reserved a subset of population from each cohort as the reference population for the clustering model, that had faster gait speeds (FG).

This analysis was approved by the institutional review board of the Albert Einstein College of Medicine, NY. All participating cohorts have received approval from their local ethics committees.

### 2.2. Study Measures

Data from all the cohorts included population demographics. To harmonize this data and address cohort variability, we followed a standardized process for the preparation of each modality of data.

**Gait Speeds:** Gait speed (cm/s) was available in all the studies. More detailed description of gait speed measurements and quantification were presented in previous studies ^5,32,33^. Gait speed was normalized within each study, then the individuals with the normalized gait speed greater than 1 standard deviation above the mean of the study population were labelled as FG. The study population was the remaining individuals with gait speeds less than 1 SD above population mean (normal and slower gait speeds). The FG participants from all the studies were used as the reference population for the HYDRA clustering algorithm.

**Volumetric MRI:** MRI measures of all the individuals across all the cohorts were available, which were collected at the respective study sites and harmonized at a single site using the FreeSurfer pipeline, using the standard parcellation and correction methods, as described elsewhere ^13^. For this study, we used the volumetric measures of 41 brain regions (Supplementary Table 1), both the hemispheres added where applicable, and total Intra Cranial Volume (tICV). To account for the individual differences in tICV, within each cohort, the volume of each region (VR) was normalized according to the mean tICV of the population within that cohort. The adjusted volume (VRa) of a brain region of an individual was calculated as

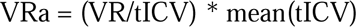

**Neuropsychological evaluations:** For the post-hoc analyses of objective cognitive scores of different subgroups obtained in our analyses, we used available neuropsychological scores from different studies, normalized per study, wherever applicable. The following were the neuropsychological tests that were used :

- Trail Making Test, Part B (TMT-B) ^34^; available in CCMA, LonGenity, EAS, NCGG-SGS (evaluated using tablet version ^35^), KES and GAIT
- Stroop Color Word test ^36^, consisting of subtests measuring time required to name the colors seen (COLOR), read the given words (WORD) and name the color of the printed word (COLOR_WORD); we used Interference (Stroop-INT) which is the difference between the third subtask and the first task (COLOR_WORD - COLOR), available in EAS, TASCOG and GAIT
- Free and Cued Selective Reminding Test (FCSRT) ^37^, a recall test that uses either words or images. Scores includes the sum of free recall (FR) alone (range 0-48) and combined with cued recall as total recall (TR), and the sum of FR and TR (FR96: range 0-96); available in CCMA, EAS and LonGenity

**Diagnosis of MCR:** MCR is characterized by slow gait accompanied by cognitive complaints. In our study, slow gait was defined as gait speed lesser than one standard deviation below the age and sex-specific means in each cohort ^38^. Gait speed in older adults from different cohorts and countries can vary due to differences in factors such as height, sex, race and ethnicity, education, and socioeconomic status.^39,40^ Hence, using a universal single gait speed cut score, although used in some prior studies to define normal or abnormal performance, is problematic as it introduces greater heterogeneity between different populations. Different cognitive complaint questionnaires were used across the 7 cohorts, sometimes with difference in their versions. For our study, subjective cognitive complaint was determined using the memory item from Geriatric Depression Scale (GDS) uniformly in all cohorts^41^ or the Instrumental Activities of Daily Living (I-ADL) ^42^, where available in cohorts CCMA and KES.

### 2.3. Analytic Approach

Figure 1 depicts the overall analytic pipeline of our study. The primary goal of this study was to identify subgroups with neuroanatomical heterogeneity among individuals with slower gait speeds using a machine learning clustering algorithm. There were as many as 41 brain region volumes available through preprocessing of structural MRIs via the automated FreeSurfer pipeline. Some of the brain region volumes are highly correlated with each other and using all of them together in a clustering model may lead to skewness and bias in the clustering results and making them less representative of the true underlying patterns in the data. Moreover, in high-dimensional spaces with many correlated features, there’s an increased risk of finding weak correlations that may result in misleading subgroup formations. Therefore, we used Factor Analyses (FA) to reduce the number of input variables to the clustering model that results in heterogeneous subgroups of population.

**Figure 1:**
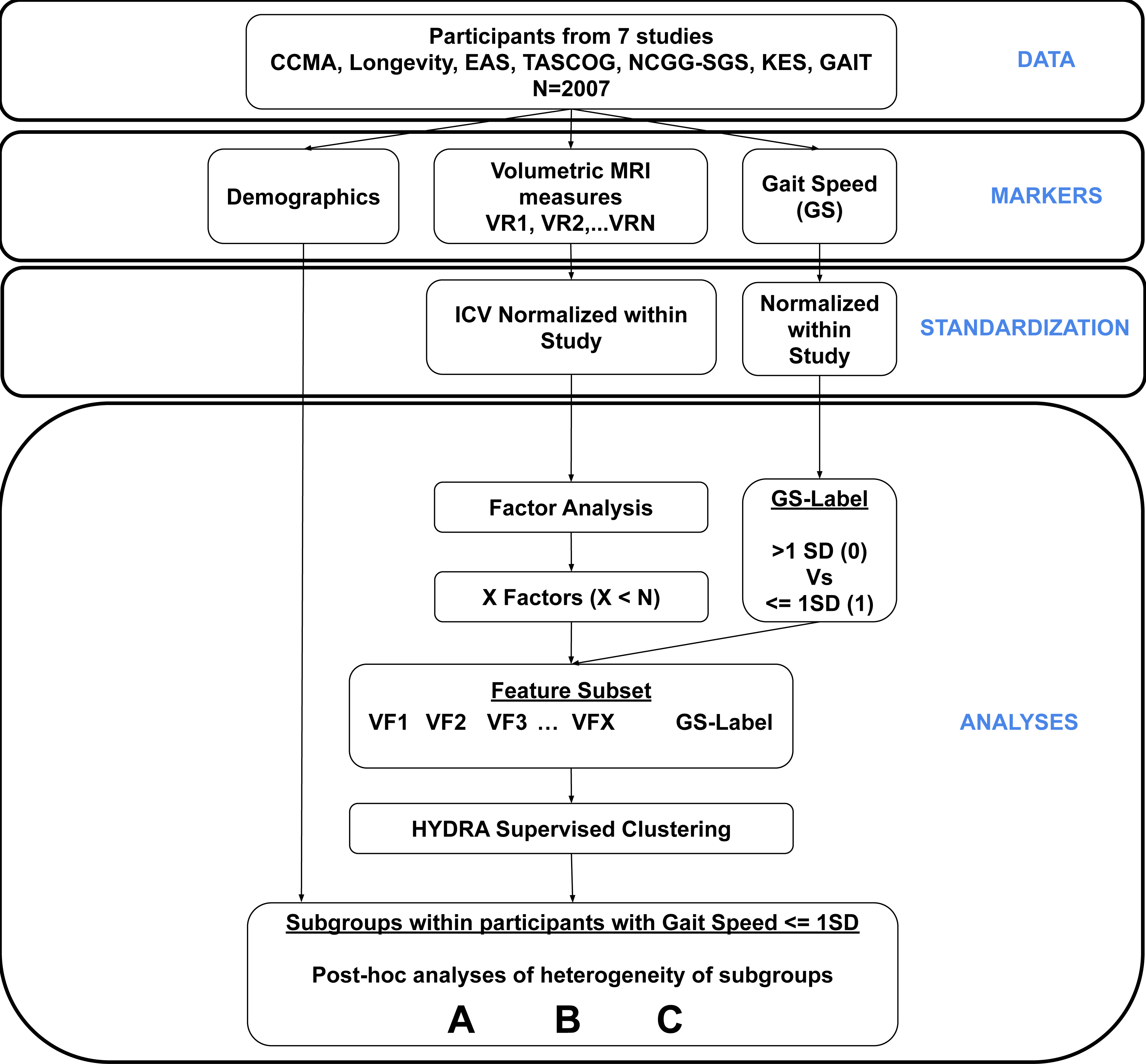
Study Plan. Plan of our study. CCMA: Central Control of Mobility in Aging Study in the US, the LonGenity study in the US, EAS : Einstein Aging Study in the US, TASCOG : Tasmanian Study of Cognition and Gait in Australia, NCGG-SGS: the National Center for Geriatrics and Gerontology–Study of Geriatric Syndromes in Japan, KES: Kerala-Einstein Study in India, and GAIT: the Gait and Alzheimer’s Interactions Tracking study; MRI: Magnetic Resonance Imaging; ICV: Intracranial Volume; SD: Standard Deviation;

Factor Analyses: For the Factor Analyses, we used maximum likelihood estimation method and varimax rotation with the 41 regions available as input variables to obtain a substantially smaller number of latent factors. The relationship between each variable (region) and the latent factors is described by a matrix of weights, or factor loadings, generated from an FA model. We chose one variable within each factor that had the highest loading to make the feature set (Regions_Factor_) for the subsequent clustering model. Therefore, before performing factor analysis, we thoroughly evaluated the “factorability” of the combined all-cohort dataset. Bartlett’s test of sphericity ^43^, which checks whether or not the observed variables intercorrelate, returned a significant value of χ^2^ = 48722.9, p<0.0001. If the test found statistically insignificant, the dataset would not have been suitable for factor analysis. We also performed the Kaiser-Meyer-Olkin (KMO) test which assesses the suitability of data for FA by measuring the sampling adequacy for each variable using the proportion of variance ^44^. The overall KMO value of our dataset was 0.94 indicating a more than adequate sampling in the data (KMO value below 0.6 is considered insufficient to proceed with FA). Thus, we proceeded with the Factor Analyses of the dataset with the available 41 volumetric MRI variables. We must specify the number of factors desired to the Factor Analyzer method being used. We performed an exploratory FA with several possible number of factors up to 25 and chose the number of factors for the model using the Kaiser criterion ^44^, based on the eigenvalues of the factors greater than 1, which represent the amount of variance explained by each factor.

Weakly-Supervised Clustering: Traditional clustering methods in machine learning are referred as ‘unsupervised’ clustering where only specific measures from the desired population are used as input for the model that results in the subgroups (clusters) of that population. The challenge with unsupervised clustering approaches is that they are not capable of accounting for ‘healthy state’ or ‘disease state’, and clustering is only performed on the specified measure (e.g., imaging biomarkers). To mitigate this problem and to obtain the subgroups in our study population relative to the FG group (i.e. *healthy state* in our population), we used a novel clustering variant, a weakly-supervised algorithm called HYDRA (Heterogeneity through Discriminative Analysis), that also considers the characteristics of population in FG group with faster gait speeds as reference, such that the population to be clustered can be simultaneously separated from the controls (FG) while quantifying the heterogeneity within the population through their association to the sub-classifiers that separate them from the reference group ^18^. As input features, we used only those regions that had the highest loading within each factor in the result of FA in the previous step. Age and sex were added as covariates to the clustering algorithm.

Statistical Analyses: The descriptive characteristics of the entire population, each cohort, and subgroups resulted from clustering were examined. The demographic variables such as age, sex, education, and ethnicity were included in the descriptive characteristics. An Analysis of Variance (ANOVA) was conducted for the quantitative demographics such as age and education. For categorical variables such as ethnicity and sex, Fisher’s exact test was performed. For the volumetric MRI measures, gait speeds and cognitive scores, an ANCOVA analysis was performed, which included age and sex as covariates to identify differences across the subgroups. For the MRI measures, we first compared a set of regions known to be associated with MCR based on previous studies ^13^. We also compared the volumes of regions used as input for the clustering model. Similar analyses were performed for gait speeds and cognitive scores across the subgroups. In case of significant differences found in the ANCOVA analyses, pairwise comparisons were performed accounted by a post-hoc Tukey’s honest significant difference (HSD) test (p < 0.05).

All the data preprocessing, Factor Analyses and HYDRA clustering were performed using Python with scikit-learn library for FA and mlni package for HYDRA clustering ^18^. The statistical analyses were conducted using the R statistical software, version 4.2.2 for MacOS.

## 3. Results

### Population Characteristics

A total of 1,987 participants were part of the final clustering model after excluding those with missing data or outliers in their volumetric MRI measures (mean age, 71.73±6.87 years; 47.7% female). There were 294 participants in the reference FG group (mean age, 69.30±5.52 years; 47.3% female) and 1693 participants in the remaining study population (mean age, 72.15±6.99 years; 47.8% female). The FG group had younger population compared to the study population (p<0.001, effect size of -0.52). Both the groups differed in the mean years of education of the population (p=0.03, effect size of 0.14). There were no sex-differences found between the groups. There was no significant difference found in both the groups in terms of cohort membership. The reference group FG differed from the study population in the proportion of participants with MCR diagnosis, as defined by ‘slow gait’, depending on threshold specific to the cohort, and subjective cognitive complaint as determined by either GDS or ADL (3.3% in the FG group, 13.3% in the study population, p<0.001) (see **Table 1**).

**Table 1:** Study Characteristics.

FG: Fast Gait, a subgroup of participants with gait speed greater than 1 standard deviation above mean; Groups A,B&C : gait speed lower than 1 SD above cohort specific mean; Subgroups obtained by the clustering model.
|  | **All participants** | **FG** | **Group A** | **Group B** | **Group C** | **P value** |
| --- | --- | --- | --- | --- | --- | --- |
| **N** | 1987 | 294 | 585 | 455 | 653 |  |
| **Age, mean (SD), y** | 71.73(±6.87) | 69.30(±5.52) | 72.07(±6.80) | 70.94(±6.77) | 73.06(±7.18) | <0.001 |
| **Sex, Female (%)** | 948(47.7%) | 139(47.3%) | 277(47.4%) | 229(50.3%) | 303(46.4%) | 0.625 |
| **Education, mean (SD), y<sup>#</sup>** | 12.30(±3.50) | 12.73(±3.70) | 12.27(±3.51) | 12.39(±3.22) | 12.08(±3.56) | 0.08 |
| **Race/Ethnicity, N(%)** |  |  |  |  |  | 0.016 |
| **Asian** | 1146(57.7%) | 176(59.9%) | 329(56.2%) | 282(62.0%) | 359(55.0%) |  |
| **White** | 769(38.7%) | 112(38.1%) | 225(38.5%) | 160(35.2%) | 272(41.7%) |  |
| **Other\*** | 59(3.6%) | 2(2.0%) | 27(5.3%) | 9(2.8%) | 21(3.3%) |  |
| **Gait Speed, cm/s** | 108.42(±22.85) | 141.21(±11.52) | 104.91(±17.05) | 103.79(±18.28) | 100.04(±21.34) | <0.001 |
| **MCR, Positive\*\*, N(%)** | 117(6.6%,N=1778) | - | 38(7.4%,N=515) | 26(6.4%,N=405) | 53(9%,N=588) | <0.001 |

p-values reported for significant differences between FG and Subgroups;
<sup>#</sup> Education was not available for the GAIT cohort.
\*Other in race/ethnicity included primarily Black (3.0% of total population), Hispanic Black, Hispanic White and others.
\*\*MCR positive is defined as slow gait (as defined by a cohort specific threshold) and any cognitive complaint in Geriatric Depression Scale (GDS) or Assisted Daily Living (ADL) questionnaire.

### Factor Analyses

We found the number of factors to be 8 and conducted FA with the same. **Supplementary Table 1** summarizes the factor loadings of each variable in all the 8 factors. For simplicity, we presented only those variables which had loadings of at least 0.4 in any factor. We chose one variable (region) from each factor that had the highest loading as the input for clustering model. The eight regions that were chosen to be the input features for the clustering model, Regions_Factor_, were the following - Accumbensarea, Caudate, Banks of the Superior Temporal Sulcus (banksSTS), Caudal Anterior Cingulate, Entorhinal Cortex, Pars Triangularis, Pericalcarine, and Precentral Gyrus.

### HYDRA Clustering Analyses

We obtained 3 subgroups within the study population from HYDRA, with an adjusted rand index (ARI) of 0.1. We performed statistical analyses to assess the differences across the 3 subgroups, Group A, Group B and Group C, between each of them as well as with respect to the FG group. **Table 1** includes descriptive characteristics of the population in each subgroup obtained. Of the 1693 individuals, 585 were grouped under Group A (mean age, 72.07±6.80 years; 47.4% female), 455 were grouped under Group B (mean age±SD, 70.94±6.77 years; 50.3% female), and 653 under Group C (mean age, 73.06±7.18 years; 46.4% female). The mean gait speed (±SD) in Group A was 104.91(±17.05), 103.79(±18.28) in Group B and 100.04(±21.34) in Group C.

### Heterogeneity in Brain Patterns in the MCR Consortium participants

Post-hoc pairwise group comparisons revealed significant differences across the subgroups for most of the regions (**Table 2 and** Figure 2**, Factors**). Among all the subgroups, Group A had the highest volumes of Accumbensarea (mean z-score of 0.481±0.140), followed by the group FG (0.434±0.126), Group B (0.425 ±0.106) and Group C (0.329±0.106). Group A had the highest volumes of Caudal Anterior Cingulate (mean z-score of 0.513±0.117), followed by the Group B (0.478±0.113), FG (0.465±0.126), and Group C (0.392±0.118). Except the Groups FG & B, all the pairs of groups differed significantly in their volumes of Accumbensarea and Caudal Anterior Cingulate (p<0.0001). All the pairs of subgroups differed significantly in their volumes of banksSTS, parstriangularis, pericalcarine and precentral gyrus. While Group A had the highest volume of banksSTS and precentral gyrus, Group B had the highest volume of parstriangularis and pericalcarine. Group B & FG did not differ significantly in their precentral gyrus volumes.

**Figure 2:**
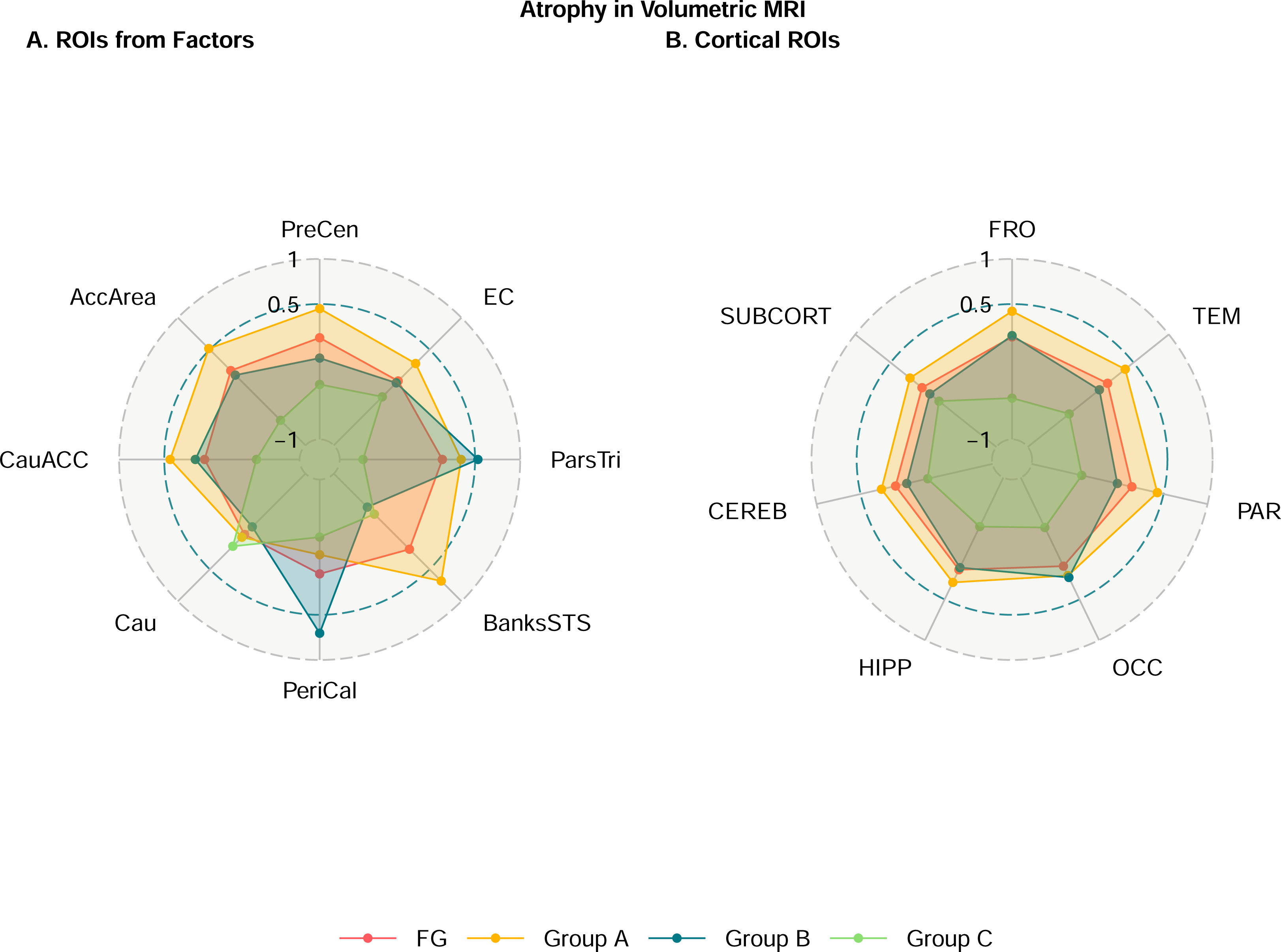
Subgroup differences in MRI Volumes by regions. Group differences in MRI Volumes.: PreCen: Precentral Gyrus, EC=Entorhinal Cortex, ParsTri=Parstriangularis, BanksSTS=Banks of Superior Temporal Sulcus, PeriCal=Pericalcarine, Cau=Caudate Nucleus, CauACC=Caudal Anterior Cingulate, AccArea=Nucleus Accumbens, FRO=Frontal Cortex, PAR=Parietal Cortex, TEM=Temporal Lobe, OCC=Occipital Lobe, HIPP=Hippocampus, CEREB: Cerebellar Cortex, SUBCORT=Subcortical regions (Basal Ganglia nuclei, Amygdala, Thalamus)

**Table 2:** Subgroup differences in Brain Region volumes - Factors.

| Measure | FG | Group A | Group B | Group C | Pairwise Comparisons (Tukey) |  |  |  |  |  |
| --- | --- | --- | --- | --- | --- | --- | --- | --- | --- | --- |
|  |  |  |  |  | Group A - FG | Group B - FG | Group C - FG | Group A - Group B | Group C - Group A | Group C - Group B |
| **Regions**<sup>Factor</sup> |  |  |  |  |  |  |  |  |  |  |
| Accumbensarea | 0.434<br>(±0.126) | 0.481<br>(±0.140) | 0.425<br>(±0.106) | 0.329<br>(±0.106) | *** | 0.92 | *** | *** | *** | *** |
| Caudate | 0.277<br>(±0.121) | 0.283<br>(±0.108) | 0.262<br>(±0.102) | 0.301<br>(±0.151) | 1 | 0.22 | 0.14 | 0.05 | 0.08 | *** |
| bankssts | 0.488<br>(±0.129) | 0.551<br>(±0.107) | 0.404<br>(±0.101) | 0.418<br>(±0.110) | *** | *** | *** | *** | *** | *** |
| caudalanteriorcingulate | 0.465<br>(±0.126) | 0.513<br>(±0.117) | 0.478<br>(±0.113) | 0.392<br>(±0.118) | *** | 0.34 | *** | *** | *** | *** |
| entorhinal | 0.508<br>(±0.097) | 0.536<br>(±0.096) | 0.506<br>(±0.090) | 0.484<br>(±0.105) | *** | 0.96 | 0.55 | *** | *** | 0.14 |
| parstriangularis | 0.576<br>(±0.106) | 0.599<br>(±0.103) | 0.620<br>(±0.099) | 0.476<br>(±0.080) | *** | *** | *** | 0.02* | *** | *** |
| pericalcarine | 0.436<br>(±0.168) | 0.403<br>(±0.134) | 0.540<br>(±0.146) | 0.372<br>(±0.140) | 0.04* | *** | *** | *** | 0.003** | *** |
| precentral | 0.528<br>(±0.109) | 0.565<br>(±0.106) | 0.502<br>(±0.108) | 0.468<br>(±0.113) | *** | 0.06 | *** | *** | *** | 0.008** |

**Regions**<sup>Factor</sup> Brain regions chosen from each of the factors from Factor Analyses, with one region with the highest loading within a factor. Precentral: Precentral Gyrus, entorhinal: Entorhinal Cortex, BanksSTS=Banks of Superior Temporal Sulcus, caudate: Caudate Nucleus \*\*\* P-values < 0.001

Next, we combined the volumes of brain regions into respective cortical, subcortical and other regions that were generally implied in MCR and dementia - C/S ROIs – Frontal, Temporal, Parietal, Occipital, Hippocampus, Cerebellum and Subcortical (Basal Ganglia nuclei, Amygdala, and Thalamus). We repeated the ANCOVA analyses and post-hoc pairwise comparisons to examine the differences across the subgroups. Differences in the C/S ROIs between the subgroups is reported in **Table 3**; Figure 2**, Cortical**). Group B did not show any significant differences in the volumes of C/S ROIs compared to that of FG, except in the Occipital lobe. Group A showed significant differences in all the C/S ROIs volumes compared to those of Group B, except in the Occipital lobe. Group A had the highest volumes across all the C/S ROIs while Group C had the least. Group C showed significant differences in the all the C/S ROIs volumes compared to those of FG, except in the Subcortical regions. Group C showed significant differences in the all the C/S ROIs volumes compared to those of Group B, except in the Cerebellum and Subcortical regions. Finally, Group C and Group A differed across all the C/S ROIs volumes.

**Table 3:** Subgroup differences in Brain Region volumes – Cortical and Subcortical Regions.

| Measure | FG | Group A | Group B | Group C | Pairwise Comparisons (Tukey) |  |  |  |  |  |
| --- | --- | --- | --- | --- | --- | --- | --- | --- | --- | --- |
|  |  |  |  |  | Group A - FG | Group B - FG | Group C - FG | Group A - Group B | Group C - Group A | Group C - Group B |
| **C/S ROIs** |  |  |  |  |  |  |  |  |  |  |
| **Frontal** | 0.300<br>(±0.030) | 0.309<br>(±0.028) | 0.300<br>(±0.027) | 0.279<br>(±0.030) | *** | 0.47 | *** | *** | *** | *** |
| **Temporal** | 0.320<br>(±0.041) | 0.330<br>(±0.038) | 0.315<br>(±0.041) | 0.296<br>(±0.042) | *** | 0.99 | *** | *** | *** | *** |
| **Parietal** | 0.312<br>(±0.032) | 0.322<br>(±0.031) | 0.306<br>(±0.029) | 0.293<br>(±0.034) | *** | 0.47 | *** | *** | *** | *** |
| **Occipital** | 0.354<br>(±0.045) | 0.359<br>(±0.042) | 0.360<br>(±0.040) | 0.333<br>(±0.043) | *** | 0.002** | *** | 0.78 | *** | *** |
| **Hippocampus** | 0.371<br>(±0.058) | 0.381<br>(±0.057) | 0.370<br>(±0.061) | 0.339<br>(±0.059) | *** | 0.71 | *** | *** | *** | *** |
| **Cerebellum** | 0.404<br>(±0.046) | 0.411<br>(±0.047) | 0.398<br>(±0.046) | 0.386<br>(±0.049) | *** | 0.79 | 0.01* | *** | *** | 0.09 |
| **Subcortical<sup>§</sup>** | 0.321<br>(±0.033) | 0.327<br>(±0.030) | 0.317<br>(±0.033) | 0.312<br>(±0.038) | *** | 0.86 | 0.66 | *** | *** | 0.98 |

**C/S ROIs :** Brain regions grouped into major cortical regions. <sup>§</sup>**Subcortical :** Includes Basal Ganglia nuclei, Amygdala and Thalamus.
\*\*\* P-values < 0.001

### Heterogeneity in Gait Speeds and Cognition in the MCR Consortium participants

ANCOVA analyses showed that the subgroups differed in their gait speeds. A post-hoc analysis was performed to evaluate subgroups differences. While Group B did not show a significant difference with either Group A or Group C, the mean gait speed of Group A was higher than that of Group C (-0.192±0.739 vs -0.364±0.864, p=0.003). In the ANCOVA analyses with each cognitive score as the outcome, all subgroups differed in their TMT-B and FR96 scores but not in their Stroop-INT scores. For TMT-B, the FG group had better scores (mean±SD: -0.354±0.892) compared to Group B (-0.066±0.906, p=0.03), Group A (-0.044±0.905, p=0.05), and Group C (0.174±1.153, p<0.001). Within the subgroups, only Groups A and C showed a significant difference (p=0.02). With respect to FR96 scores, the FG group had higher scores (0.599±0.716) compared to Group C (-0.125±1.136, p=0.02). Group A also had higher scores (0.389±0.778) compared to Group C (p=0.01).The groups did not show significant differences in their Stroop-Int scores. **Table 4** and Figure 3 summarize the ANCOVA and post-hoc pairwise comparison results for gait speed, and cognitive scores across the subgroups.

**Figure 3:**
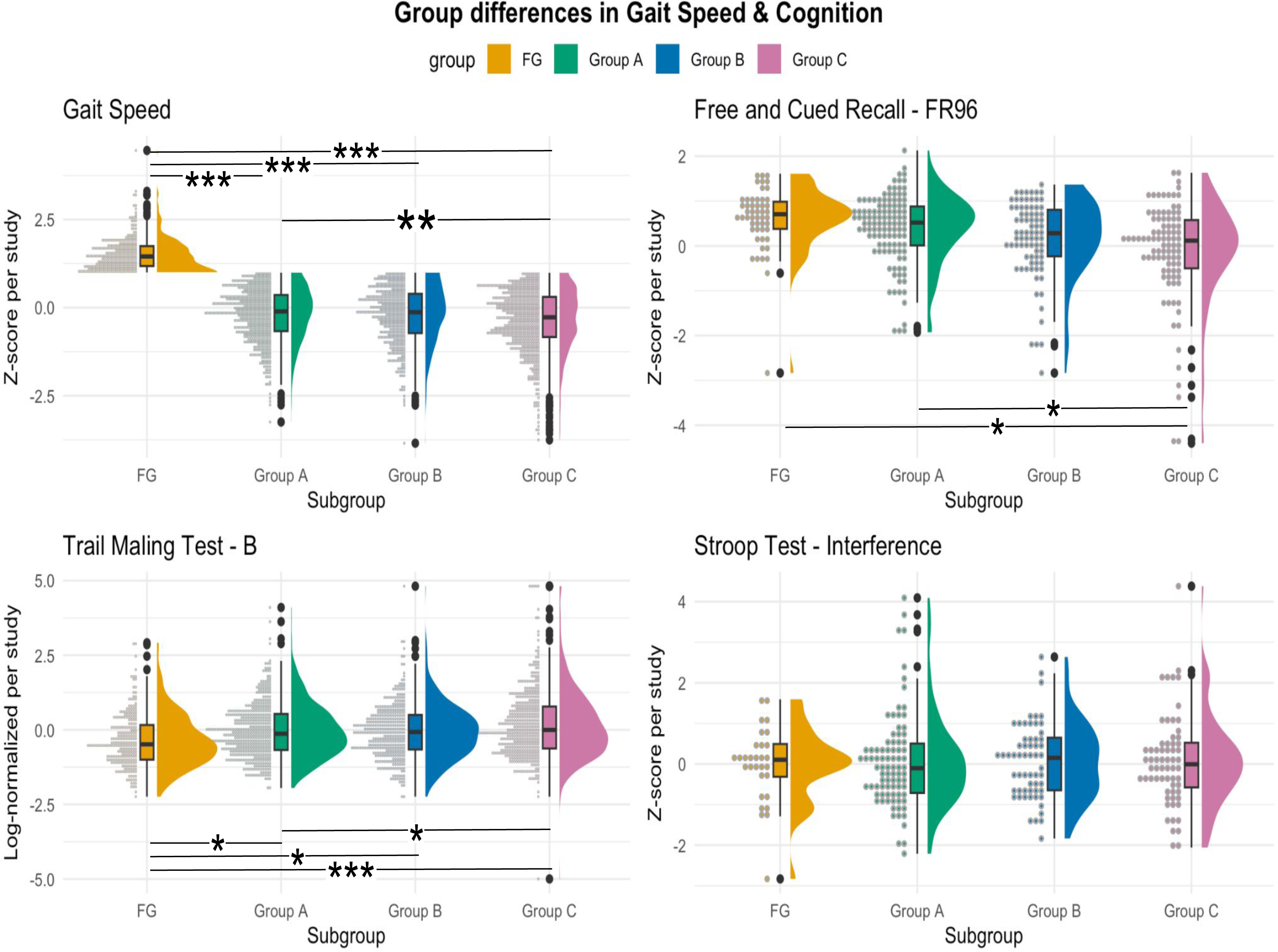
Subgroup differences in MRI Volumes by regions. Group differences in Gait speed and Cognition. TMT-B : Trail Making Test, Part B, log-transformed, z-score normalized, available in CCMA, LonGenity, EAS, NCGG-SGS, Kes and GAIT. FR96: Free and Cued Selective Reminding Test (FCSRT), a recall test that uses either words or images. Scores includes the sum of free recall (FR) alone (range 0-48) and combined with cued recall as total recall (TR), FR96: the sum of FR and TR (range 0-96); available in CCMA, EAS and LonGenity; Stroop-INT : Stroop Color Word test, consisting of subtests measuring time required to name the colors seen (COLOR), read the given words (WORD) and name the color of the printed word (COLOR_WORD); Stroop-INT: the difference between the third subtask and the first task (COLOR_WORD - COLOR), available in EAS, TASCOG and GAIT.

**Table 4:** Pairwise Subgroup Differences in Gait speed and Cognition.

| Measure | FG | Group A | Group B | Group C | P-value | Group A - FG | Group B - FG | Group C - FG | Group A - Group B | Group C - Group A | Group C - Group B |
| --- | --- | --- | --- | --- | --- | --- | --- | --- | --- | --- | --- |
| Gait Speed (Z-score) | 1.531<br>(±0.466) | -<br>0.192<br>(±0.739) | -0.223<br>(±0.793) | -0.364<br>(±0.864) | *** | *** | *** | *** | 0.58 | 0.003** | 0.22 |
| **Cognition** |  |  |  |  |  |  |  |  |  |  |  |
| TMT-B | -0.354<br>(±0.892) | -<br>0.044<br>(±0.905) | -0.066<br>(±0.906) | 0.174<br>(±1.153) | *** | 0.05* | 0.03* | *** | 0.98 | 0.02* | 0.09 |
| FR96 | 0.599<br>(±0.716) | = | 0.148<br>(±0.871) | -0.125<br>(±1.136) | 0.005** | 0.95 | 0.23 | 0.02* | 0.31 | 0.01* | 0.61 |
| Stroop-INT | -0.025<br>(±0.918) | 0.069<br>(±1.175) | 0.061<br>(±0.923) | 0.076<br>(±1.122) | 0.99 | - | - | - | - | - | - |

\*\*\* P-values < 0.0001

**Table 5:** MCR Consortium Participant Subgroup Characteristics.

FG: Population with fast gait, gait speed greater than 1SD above population-mean; **Groups A,B&C:** result of HYDRA Clustering of the remaining population (gait speed slower than 1 SD above population-mean); **P-value:** Significance $Pr(>F)$ in ANCOVA corrected for age and sex; **Pairwise comparisons:** Multiple Comparisons of Means with Tukey Contrasts.
| Group | Gait speed, Imaging, and Cognition |
| --- | --- |
| **Fast Gait (FG)**<br>**Gait Speed > mean+1SD** | Reference group, used by the clustering algorithm<br>Best scores in FCSRT and TMT-B, No cortical or subcortical atrophy |
| **Study Population**<br>**Gait Speed < mean+1SD** |  |
| **Group A** | Fastest gait, largest volumes across almost all ROIs |
| **Group B** | Intermediate gait speed, similar cortical volumes as reference group |
| **Group C** | Slowest gait, lowest cortical volumes, highest prevalence of MCR |

## 4. Discussion

The MCR Consortium comprises over 2,000 MRIs from older adults without dementia from multiple locations across different continents. It provides an invaluable repository of neuroimaging and other measures to further our understanding of MCR as a crucial risk factor for dementia. Using a novel data-driven machine learning approach, we focused on exploring the heterogeneity in the population with fast and normative gait speeds (slower than 1 SD above the population mean), while accounting for the distinct neuroanatomical patterns that separate each subgroup from the population with fast gait speeds. Among the participants without dementia and gait speeds in the normal to slow range, we found three subgroups which significantly differed in the patterns of brain regional volumes, gait speeds, and cognitive performance.

Group A had slower gait speeds compared to the reference fast gait speed (FG) group and largest volumes of cortical and subcortical regions. Group B had intermediate gait speeds and cortical volumes similar to the reference group. Group C had the slowest gait speeds, the smallest overall regional brain volumes in most brain regions and the highest prevalence of MCR. The subgroups also differed in their performance in Trail Making Test (Part B) and Free & Cued Selective Reminder Test.

Abnormal gait has been shown to be a reliable predictor of non-Alzheimer dementia ^45,46^. Since slowing of gait speed precedes cognitive decline^32,33^, understanding the heterogeneity related to gait speed patterns can be very valuable in characterizing predementia syndromes such as MCR. MCR, which is characterized by slow gait and subjective cognitive impairment, depends on varying thresholds to determine slow gait ranging from 44.4-101.9 cm/s among men and 36.9-97.4 cm/s among women as observed across different study locations ^5^. In our analyses, instead of studying abnormal gait using fixed thresholds, we considered a broader group of older population without dementia with gait speeds that are fast or normative (less than 1 SD above population mean). We were interested in studying neuroanatomical patterns within this population in relation to their gait speed and in exploring possible differences in their cognition. As hypothesized, we found homogenous subgroups that showed significant differences in their mean gait speeds accompanied by corresponding distinct neuroanatomical patterns as shown by differences in their brain region volumes. The subgroup with the slowest mean gait speed (Group C) had the least volumes across the prefrontal, temporal, and parietal cortex regions, hippocampus and cerebellar cortex than any other subgroups, resembling a pathological pattern of an MCR subtype accompanied by olfactory dysfunction that is more likely to be associated with Alzheimer and Lewy body dementias ^47^. Our results were consistent with the previous MCR studies that showed higher association of lower cortical gray matter volumes and total hippocampal volumes with incident MCR ^8,9,13^. Similar results were observed across the regions that were used in the clustering model, except for the Caudate nucleus, for which no significant difference was found across any of the groups. Interestingly, Group A, which had a slower mean gait speed compared to the reference fast gait group, had higher brain region volumes across prefrontal, hippocampal, temporal, parietal regions. We could not find a specific neuroanatomical pattern that could explain why Group A may have slower gait speeds than the reference group FG while having greater brain region volumes. This finding might be explainable by other comorbidities. However, due to the lack of detailed data on these comorbidities, we were not able to study them in the context of the current study.

Prior studies indicate that MCR syndrome is associated with worse performance in different cognitive domains such as attention and memory, as well as global cognition ^48^. In a study of 314 nondemented older adults ^49^, it was shown that different MCR subtypes show differences in cognitive profiles, with different gait attributes such as speed, stride length and swing being differently associated with global cognition and memory. In this study, we evaluated possible subgroup differences in their cognitive performance as measured by different tests. We compared the performances of the individuals across all the subgroups in TMT-B, FCSRT (as measured by FR96) and Stroop Task - Interference. All the subgroups showed worse performance in TMT-B and FCSRT compared to that of the faster gait speed group. All the subgroup pairs, except Groups A and B, differed in their TMT-B scores, underscoring the previously established links between executive function and gait speed, accompanied by cognitive complaints^8,15^. Whereas only the slowest gait subgroup, Group C, differed in its mean FR96 scores compared to FG and Group A. None of the subgroups differed in their cued recall scores and Stroop- Interference scores, which is likely attributable to the fact that the study populations were all dementia-free at baseline visit. Our findings underscore various results from previous studies that highlighted the negative association between gait speed and impairment in executive function and global cognition ^48,50,51^.

We used the data from a large multi-center consortium of studies involving community-dwelling non-demented adults. However, a few limitations should be noted. Due to cross-sectional analyses, it is not possible to establish any causal relationships between identified structural brain changes among groups and clinical outcomes. A future direction for us is to study if these subgroups that differ in their neuroanatomic and gait speed patterns, would show different trajectories in their gait speed slowdown or cognitive decline. Previous studies that both cortical volume and thickness correlate with cognitive function and neurodegeneration^52^, while particularly in the case of MCR it was shown that cortical thickness detected neurodegeneration easier than cortical volumes^13^. However, it was shown that cortical volume can be more sensitive to detecting changes in brain structure associated with aging and neurodegenerative processes^53^ and numerous studies have successfully used cortical volumes in machine learning models for various neurological conditions^54,55^. Therefore, we used cortical volumes as the inputs for our model. The effects of using cortical thickness on the resulting subgroups should be further explored. We used a novel method of combining factor analyses with weakly-supervised clustering, which might not be necessarily the most fitting approach to study the relation between gait speeds and neuroanatomical heterogeneity. However, this approach handles the problem of collinearity in brain region volumes very well. We must highlight that the results of the analyses of subgroup differences in their cognitive performance may not be generalizable since different cognitive tests and different versions have been used across the cohorts and some of the cognitive tests were not available for some cohorts. In combination with longitudinal measures and harmonized cognitive measures, novel machine learning methods such as HYDRA or the most recent generative methods ^56^ that are specialized in studying heterogeneity in imaging, have a huge potential to explore disease-subtypes and disease-staging in gait abnormalities and MCR syndrome.

The significance of our work lies in its potential to encourage further research related to previously unexplored heterogeneity within the at-risk population with slower gait speeds. By identifying distinct subgroups based on brain volume patterns as well as other biomarkers, we can potentially improve our understanding of the underlying pathophysiology associated with slow gait and its relation to MCR syndrome and subsequent progression to dementia. This approach might help paving pathways toward patient stratification at early asymptomatic stages and have implications for precision health. As the multi-site study consortiums such as MCR Imaging Consortium are invaluable to the research of aging and dementia, our methodology succeeds in handling the challenges of such complex datasets, offering a robust framework for analyzing neuroimaging data across diverse populations. Ultimately, our work contributes to the growing body of knowledge on MCR and may pave the way for more nuanced approaches to early dementia detection and prevention.

## Data Availability

All data produced in the present study are available upon reasonable request to the authors

## Acknowledgements

The authors gratefully acknowledge the contributions of the participants, families, and study staff from all the studies in the consortium that provided the data for this research. We also thank the funding agencies and institutions that have supported these studies.

## Conflicts of Interests

HMB serves as a consultant for Neural+. RBL is the Edwin S. Lowe Professor of Neurology at the Albert Einstein College of Medicine in New York. He also receives support from the Migraine Research Foundation and the National Headache Foundation and research grants from TEVA, Satsuma and Amgen. He serves on the editorial board of Neurology, senior advisor to Headache, and associate editor to Cephalalgia. He has reviewed for the NIA and NINDS, holds stock and stock options in Axon, Biohaven Holdings, CoolTech and Manistee; serves as consultant, advisory board member, or has received honoraria from: Abbvie (Allergan), American Academy of Neurology, American Headache Society, Amgen, Avanir, Axon, Axsome, Biohaven, Biovision, Boston Scientific, Dr. Reddy’s (Promius), Electrocore, Eli Lilly, eNeura Therapeutics, Equinox, GlaxoSmithKline, Grifols, Lundbeck (Alder), Manistee, Merck, Pernix, Pfizer, Satsuma, Supernus, Teva, Trigemina, Vector, Vedanta. He receives royalties from Wolff’s Headache 7th and 8th Edition, Oxford Press University, 2009, Wiley and Informa. JV serves as an Advisory Committee Member for MedRhythms, and a holds a uncompensated voluntary position at CatchU. Authors BTN, AE, KKP, EA, VGPK, SV, RB, OB, TD, HS, SM, and SA do not have any conflicts of interest to disclose.

## Funding Sources

This study was supported by NIH/NIA grants: 1R56AG057548-01, R01AG057548-01A1 and 2R01AG039330 (JV), NIH RO1 DK129320-01 2021-2026 (RB), NIA R01AG062659-01A1 (HB), NIA K23 AG063993, AG080635, AG003949, the Alzheimer’s Association (SG-24-988292 ISAVRAD); Cure Alzheimer’s Fund, the Leonard and Sylvia Marx Foundation (AE, RL), NIH/NIA 1K76AG083274 (SA). None of the funding sources had any role in the conduct of the analysis, interpretation of data and preparation of the article.

## Consent Statement

All human subjects across all the study locations provided informed consent.

## Notes

### Author Declarations

Ethics committee/IRB of the Albert Einstein College of Medicine, bronx, NY, USA gave ethical approval for this work

