## Supplementary Table 1 for "Discovering Subtypes with Imaging Signatures in the Motoric Cognitive Risk Syndrome Consortium using Weakly-Supervised Clustering"

**Title**

1300 Morris Park Avenue

Bronx, NY 10461

**Supplementary Table 1: Factor Analysis of Brain Region Volumes**

| Region | F1 | F2 | F3 | F4 | F5 | F6 | F7 | F8 | Region | F1 | F2 | F3 | F4 | F5 | F6 | F7 | F8 |
| --- | --- | --- | --- | --- | --- | --- | --- | --- | --- | --- | --- | --- | --- | --- | --- | --- | --- |
| *Accumbensarea* |  |  |  |  |  |  |  | 0.49 | lingual |  |  |  |  | 0.7 |  |  |  |
| Amygdala |  | 0.63 |  |  |  |  |  |  | medialorbitofrontal |  |  | 0.49 |  |  |  |  |  |
| *Caudate* |  |  |  |  |  | 0.77 |  |  | middletemporal |  |  |  | 0.59 |  |  |  |  |
| CerebellumCortex |  |  |  |  |  |  |  |  | paracentral | 0.6 |  |  |  |  |  |  |  |
| Hippocampus |  | 0.61 |  |  |  |  |  |  | parahippocampal |  | 0.43 |  |  |  |  |  |  |
| Pallidum |  | 0.42 |  |  |  |  |  |  | parsopercularis |  |  | 0.46 |  |  |  |  |  |
| Putamen |  |  |  |  |  | 0.76 |  |  | parsorbitalis |  |  | 0.58 |  |  |  |  |  |
| ThalamusProper |  |  |  |  |  | 0.53 |  |  | ***parstriangularis*** |  |  | 0.61 |  |  |  |  |  |
| *bankssts* |  |  |  | 0.66 |  |  |  |  | ***pericalcarine*** |  |  |  |  | 0.81 |  |  |  |
| *caudalanteriorcingulate* |  |  |  |  |  |  | 0.74 |  | postcentral | 0.61 |  |  |  |  |  |  |  |
| caudalmiddlefrontal | 0.45 |  |  |  |  |  |  |  | posteriorcingulate |  |  |  |  |  |  | 0.47 |  |
| cuneus |  |  |  |  | 0.75 |  |  |  | ***precentral*** | 0.69 |  |  |  |  |  |  |  |
| *entorhinal* |  | 0.64 |  |  |  |  |  |  | precuneus | 0.63 |  |  |  |  |  |  |  |
| frontalpole |  |  |  |  |  |  |  | 0.4 | rostralanteriorcingulate |  |  |  |  |  |  | 0.6 |  |
| fusiform |  | 0.5 |  |  |  |  |  |  | rostralmiddlefrontal |  |  | 0.53 |  |  |  |  |  |
| inferiorparietal |  |  |  | 0.58 |  |  |  |  | superiorfrontal | 0.54 |  | 0.42 |  |  |  |  |  |
| inferiortemporal |  | 0.6 |  | 0.4 |  |  |  |  | superiorparietal | 0.6 |  |  |  |  |  |  |  |
| isthmuscingulate |  |  |  |  |  |  |  |  | superiortemporal |  |  |  | 0.46 |  |  |  |  |
| lateraloccipital |  |  |  | 0.42 |  |  |  |  | supramarginal | 0.58 |  |  |  |  |  |  |  |
| lateralorbitofrontal |  |  | 0.53 |  |  |  |  |  | temporalpole |  | 0.63 |  |  |  |  |  |  |

**Note**: F1 to F8 are the 8 factors obtained by Factor Analysis. For the subsequent clustering model, one variable with the highest loading within each factor was selected: Accumbensarea, Caudate, bankssts, caudalanteriorcingulate, entorhinal, parstriangularis, pericalcarine, and precentral.
